# Alcohol Screening for Women in their Childbearing Years: What are Health Care Providers doing in Canada?

**DOI:** 10.1101/2020.03.19.20038992

**Authors:** Christina Cantin, Tejal Patel, Courtney R. Green, Kyla Kaminsky, Nicole Roberts, Jocelynn L. Cook

## Abstract

Health care providers (HCPs) have an important role in screening for alcohol use across the lifespan, particularly during the childbearing years, and providing brief intervention to yield optimal outcomes and prevent the potential teratogenic effects of prenatal alcohol exposure.

**Objective:** The purpose of this study was to describe the current alcohol screening practices of Canadian HCPs who care for pregnant women and women of childbearing age.

**Methods:** An online survey was administered in 2017 with the aim to identify current knowledge, attitudes, practices and beliefs among Canadian HCPs on screening, brief intervention and referral to treatment (SBIRT) for alcohol use for this population. The bilingual survey was disseminated by 4 national professional associations. A total of 634 interprofessional clinicians completed to the survey. Descriptive analysis was completed for the respondent’s profession and their practices related to alcohol SBIRT. Cross-tabulation analyses explored the use of different screening questionnaires.

**Results:** Most respondents reported asking about alcohol use; however, there was a low overall use of screening questionnaires for both women of childbearing age and those who are pregnant. Low screening rates may equate to missed opportunities for intervention. Low rates of brief intervention and referral were noted even in circumstances where at-risk drinking was identified, with only 16.4% of respondents intervening when pregnant women reported at-risk alcohol consumption.

**Conclusion:** Continued efforts are needed to improve alcohol screening practices among women’s HCPs across Canada. Priority areas for training include: understanding validated alcohol screening questionnaires; incorporating brief intervention into routine care; and developing local referral pathways.

## Introduction

Alcohol use during pregnancy continues to be a significant public health concern. Despite the strong evidence correlating alcohol consumption in pregnancy with adverse fetal outcomes, namely Fetal Alcohol Spectrum Disorders (FASD), there is insufficient evidence to define a safe threshold (Carson et al., 2017). Therefore, abstinence continues to be the recommendation for women who are considering a pregnancy or who are pregnant.

Alcohol consumption is a common and socially acceptable behaviour in Canadian society. According to the Public Health Agency of Canada’s Canadian maternity experiences survey, 62.4% of women of childbearing age (≥ 18 years) reported consuming alcohol three months before they were pregnant or before pregnancy recognition; 10.5% women reported consuming alcohol during pregnancy (PHAC, 2009). The unplanned pregnancy rate among *all* Canadian women is reportedly 40%, but as high as 70% in adolescents (Black et al., 2015; PHAC, 2009). This translates to a high potential rate of prenatal alcohol exposure during critical embryonic and fetal developmental stages. Therefore, health care providers (HCPs) are uniquely placed to screen for alcohol use across the lifespan, particularly during the childbearing years, and provide management strategies to mitigate harm.

Universal screening is one prevention measure that HCPs can easily incorporate into their routine practice, enabling them to identify patients who are consuming alcohol. This would allow HCPs to provide interventions and support in the preconception period (ACOG, 2011; WHO, 2014). Although some populations are at higher risk for substance use (Carson et al., 2017), the following characteristics have been identified as the most likely to be overlooked when screening for alcohol use: over 35 years of age; social drinker; highly educated; history of sexual and/or emotional abuse; and high socioeconomic status (PHAC, 2002). Furthermore, a randomized controlled trial (RCT) of 304 pregnant women and their partners identified the following factors associated with increased prenatal alcohol use: more years of education; extent of previous alcohol consumption (prior to pregnancy); and temptation to drink in social situations (Chang et al., 2005). Universal screening by definition means that *all* women receive screening for substance use regardless of their sociodemographic characteristics. Use of an appropriate and validated screening questionnaire allows for a better understanding of problematic alcohol use and can inform further referrals and treatment.

There is compelling evidence that screening for problematic alcohol use in pregnancy, followed by brief intervention (BI) for those identified to be at-risk, can decrease alcohol consumption during pregnancy (Carson et al., 2017; Chang et al., 2005; Chang, Wilkins-Haug, Berman, & Goetz, 1999; Floyd, Weber, Denny, & O’Connor, 2009), in the postnatal period (Chang et al., 2005), and during women’s childbearing years (Wilton et al., 2013). A RCT of 205 women of childbearing age receiving primary care and community-based care by physicians found a significant treatment effect in reducing both 7 day alcohol use (p<0.005) and binge drinking episodes (p<0.005) over a 48 month follow-up period (Manwell, Fleming, Mundt, Stauffacher, & Barry, 2000). Women in the experimental group, who became pregnant during the follow-up period, had the most dramatic decreases in alcohol use suggesting that BI is effective in maintaining a reduction in alcohol use.

In North America, both the American College of Obstetrics and Gynecology (ACOG) and Society of Obstetricians and Gynaecologists of Canada (SOGC) recommend universal screening for alcohol use in pregnancy (ACOG, 2011; Carson et al., 2017). Screening can take many forms. Simply asking one or two questions to identify if the woman drinks alcohol, how often or how much has been shown to effectively open a dialogue about alcohol use in pregnant women (Burd et al., 2006). Alternatively, screening can be accomplished by the use of a structured and validated questionnaire. A number of questionnaires (Table 1) have been described in the literature for this purpose (Bush, Kivlahan, McDonell, Fihn, & Bradley, 1998; Knight et al., 1999; Mayfield, McLeod, & Hall, 1974; Pokorny, Miller, & Kaplan, 1972; Russell & Bigler, 1979; Saunders, Aasland, Babor, de la Fuente, & Grant, 1993; Selzer, Vinokur, & van Rooijen, 1975; Sokol, Martier, & Ager, 1989). Among these, the T-ACE and TWEAK (Tolerance, Worry, Eye-opener, Amnesia, Kut down) were designed and validated specifically for the obstetrical population (Burns, Gray, & Smith, 2010; Sokol et al., 1989) while others, such as the AUDIT-C (Alcohol Use Disorders Identification Test – Alcohol Consumption Questionnaire) and CAGE (Cut down, Annoy, Guilt, Eye-opener), have been studied in pregnant women though they were originally developed for use in men. Currently, the evidence would suggest that the T-ACE, TWEAK and AUDIT-C screening questionnaires have the most applicability and promise for use in the pregnant population (Burns et al., 2010; Shogren, Harsell, & Heitkamp, 2017; Smith, Savory, Couves, & Burns, 2014). Accordingly, ACOG recommends use of the T-ACE questionnaire and discourages use of CAGE during pregnancy (ACOG, 2011), while the SOGC recommends using CRAFFT (Car, Relax, Alone, Forget, Family, Trouble) in adolescent pregnancies and either T-ACE or TWEAK with adult pregnant women (ACOG, 2011; Chudley et al., 2005). Premji and Semenic suggest the T-ACE scale can identify alcohol use among diverse groups of pregnant women, due to its high sensitivity and specificity (Premji & Semenic, 2009).

**Table 1:**
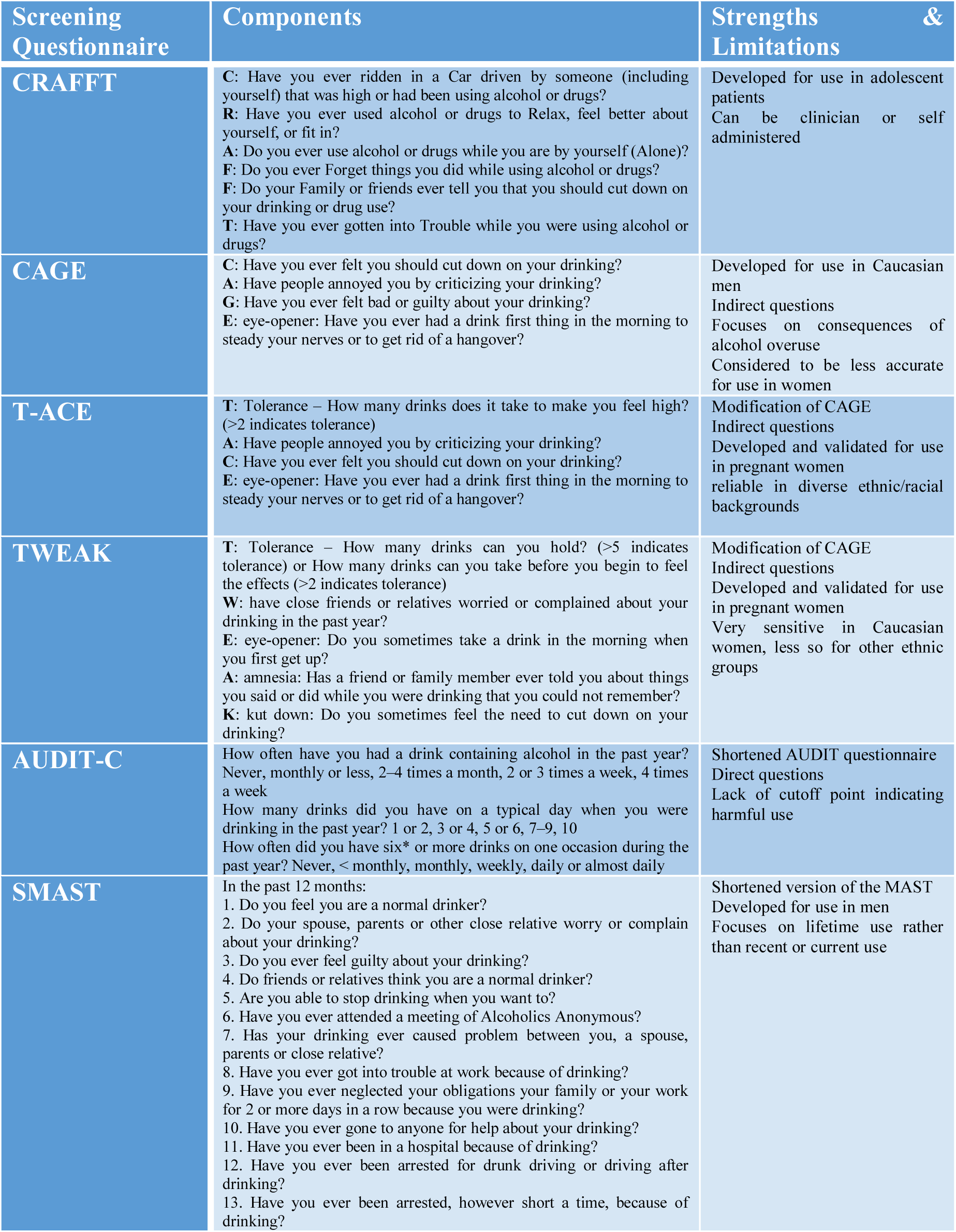
Examples of questionnaires that can be used to screen for problematic alcohol use.

Despite the recommendation, uptake of formal screening questionnaires among physicians and nurses is poor, with only 44% reporting consistent use (Davis, Carr, & La, 2008). Often cited barriers to screening include the time required to conduct the questionnaire; the need for a specific skill set and training to conduct sensitive and difficult conversations; and a perceived lack of confidence or competence among HCPs (Diekman et al., 2000; Gassman, 2003; Poole, Schmidt, Green, & Hemsing, 2016; Spandorfer, Israel, & Turner, 1999; Wangberg, 2015). The purpose of this paper is to describe the current alcohol screening practices of Canadian HCPs who care for pregnant women and women of childbearing age, to explore differences among HCP groups, and to identify priority areas for training.

## Methods

The SOGC conducted a national survey in 2017 that was based on the 2002 survey on the same topic (PHAC, 2002). An interprofessional panel was tasked with revising the 2002 survey, based on their expertise in the field and informed by a comprehensive literature review. Once the survey (consisting of 116 questions) was finalized, it was translated and administered using SurveyMonkey®. The survey was disseminated to Canadian HCPs in both English and French between October and November 2017 through the following professional association membership lists: SOGC, the College of Family Physicians of Canada (CFPC), the Canadian Association of Midwives (CAM), and the Canadian Association of Perinatal and Women’s Health Nurses (CAPWHN). Two reminder emails were sent after the initial recruitment email.

The study received ethics approval from the Ottawa Health Science Network Research Ethics Board (REB #20170407-01H). Descriptive analysis was conducted using Statistical Analysis Software (SAS) (SAS, 2019) version 9.4 reporting respondent’s profession and their practices related to alcohol screening, provision of advice, brief intervention, harm reduction, and referral to treatment. Additionally, cross-tabulations were completed by HCP group (i.e. Family Physicians, Midwives, Obstetricians/Gynaecologists (Ob/Gyns), & Nurses) to explore any differences in the types of screening questionnaires used for both pregnant and women of childbearing age.

## Results

A total of 634 HCPs completed the survey. The demographic characteristics of the respondents are summarized in Table 2. Participants from all disciplines surveyed were represented, with midwives making up a third (33.3%) of all respondents. Participants were mostly female (89.6%) and practicing in Ontario (51.5%). There was almost equal participation of HCPs by years in practice, with slightly higher rates for those in their first 5 years of practice (34.2%). One third of all respondents identified some personal experience with problematic alcohol use. Familiarity and use of the SOGC guidelines was modest.

**Table 2.**
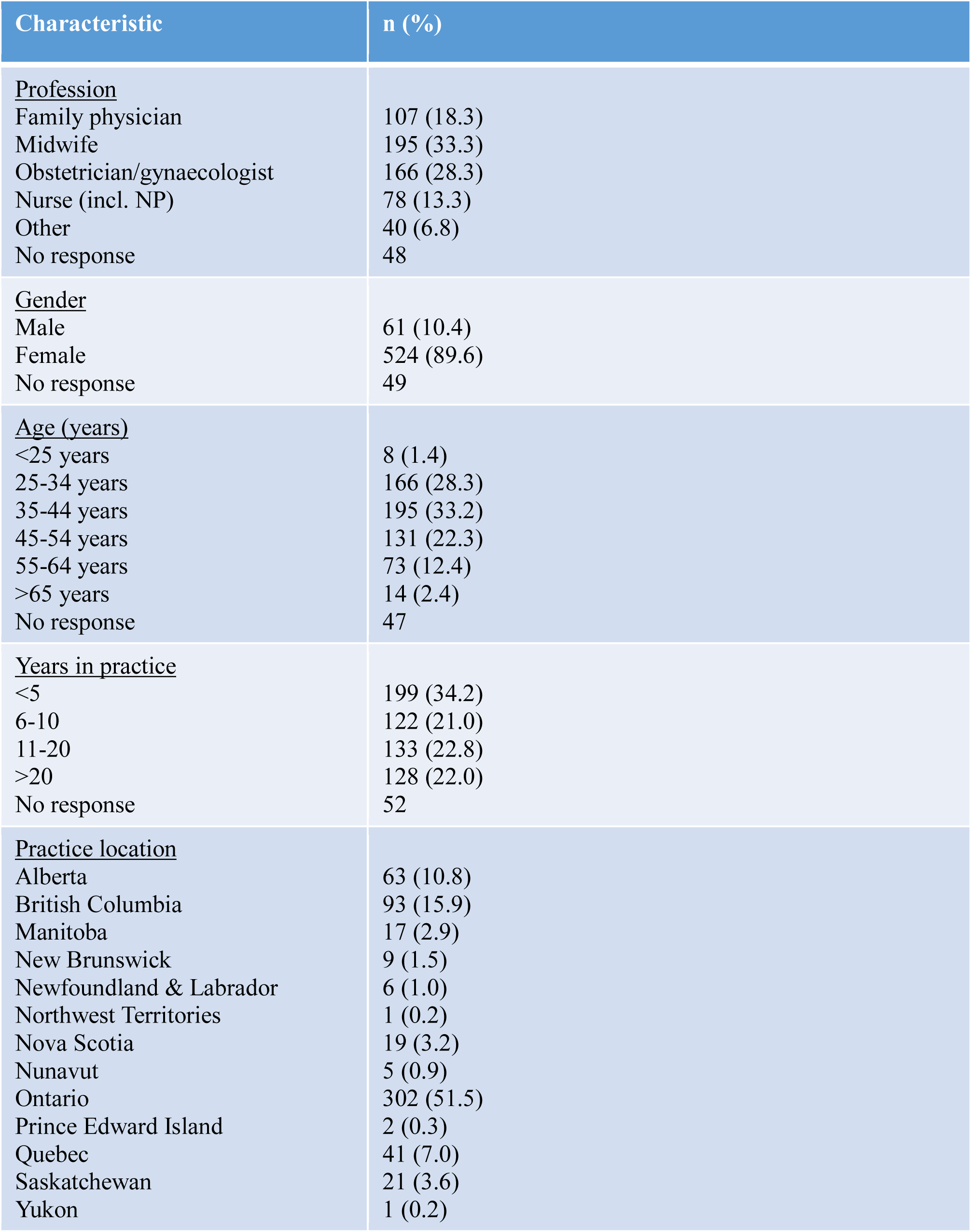
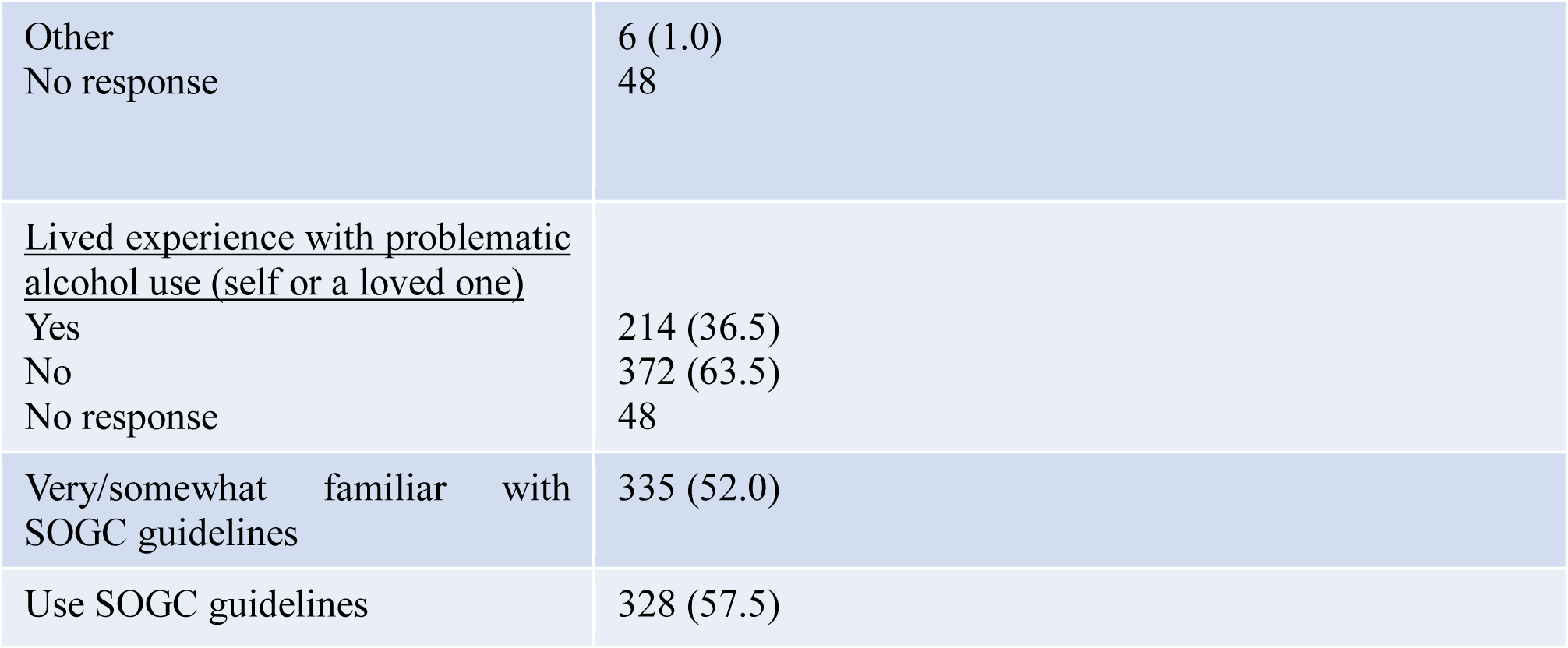
Study Participant Description.

| Characteristic | n (%) |
| --- | --- |
| <u>Profession</u> |  |
| Family physician | 107 (18.3) |
| Midwife | 195 (33.3) |
| Obstetrician/gynaecologist | 166 (28.3) |
| Nurse (incl. NP) | 78 (13.3) |
| Other | 40 (6.8) |
| No response | 48 |
| <u>Gender</u> |  |
| Male | 61 (10.4) |
| Female | 524 (89.6) |
| No response | 49 |
| <u>Age (years)</u> |  |
| <25 years | 8 (1.4) |
| 25-34 years | 166 (28.3) |
| 35-44 years | 195 (33.2) |
| 45-54 years | 131 (22.3) |
| 55-64 years | 73 (12.4) |
| >65 years | 14 (2.4) |
| No response | 47 |
| <u>Years in practice</u> |  |
| <5 | 199 (34.2) |
| 6-10 | 122 (21.0) |
| 11-20 | 133 (22.8) |
| >20 | 128 (22.0) |
| No response | 52 |
| <u>Practice location</u> |  |
| Alberta | 63 (10.8) |
| British Columbia | 93 (15.9) |
| Manitoba | 17 (2.9) |
| New Brunswick | 9 (1.5) |
| Newfoundland & Labrador | 6 (1.0) |
| Northwest Territories | 1 (0.2) |
| Nova Scotia | 19 (3.2) |
| Nunavut | 5 (0.9) |
| Ontario | 302 (51.5) |
| Prince Edward Island | 2 (0.3) |
| Quebec | 41 (7.0) |
| Saskatchewan | 21 (3.6) |
| Yukon | 1 (0.2) |
| Other<br>No response | 6 (1.0)<br>48 |
| <u>Lived experience with problematic alcohol use (self or a loved one)</u><br>Yes<br>No<br>No response | 214 (36.5)<br>372 (63.5)<br>48 |
| Very/somewhat familiar with<br>SOGC guidelines | 335 (52.0) |
| Use SOGC guidelines | 328 (57.5) |

The majority of HCPs stated that “no alcohol during pregnancy is recommended.” Interestingly, more HCPs recommended this for pregnant women than for women of childbearing age (93.1% vs 88.1%). Nearly all participants (94.9%) reported asking pregnant women about alcohol use, while only 71.9% asked women of childbearing age (Table 3). More than one-third of participants reported using an alcohol screening questionnaire (38% with pregnant women, and 42% with women of childbearing age). However, only 35% and 14% of the participants reported that they “frequently used” a screening questionnaire for pregnant women and women of childbearing age, respectively (Table 3). The majority of respondents felt that they had enough time to use a structured alcohol screening questionnaire for pregnant women (76.9%) and women of childbearing age (69.3%).

**Table 3.** Use of alcohol screening questionnaire.

| Question | Pregnant Women |  | Childbearing Women |  |
| --- | --- | --- | --- | --- |
|  | n | % | n | % |
| Do you ever use an alcohol screening questionnaire? |  |  |  |  |
| Yes | 216 | 38 | 151 | 42 |
| No | 355 | 62 | 209 | 58 |
| On average, how often do you use an alcohol screening questionnaire? |  |  |  |  |
| Frequently | 76 | 35 | 21 | 14 |
| Sometimes | 60 | 28 | 70 | 47 |
| Rarely | 79 | 37 | 59 | 39 |
| I have enough time to use a structured alcohol screening questionnaire |  |  |  |  |
| Yes | 166 | 77 | 104 | 69 |
| No | 50 | 23 | 46 | 31 |

HCPs choice of alcohol screening questionnaires is summarized in Table 4. Overall, T-ACE and CAGE were the most commonly used structured alcohol screening questionnaires (38% and 41% respectively for pregnant women; and 25% and 79% respectively for women of childbearing age). Variations in choice of screening questionnaires were revealed among different HCPs; family physicians most often used CAGE whereas midwives most often used TWEAK; Ob/Gyns were equally likely to choose T-ACE and CAGE for pregnant women, however, CAGE was preferentially used for women of childbearing age.

**Table 4.**
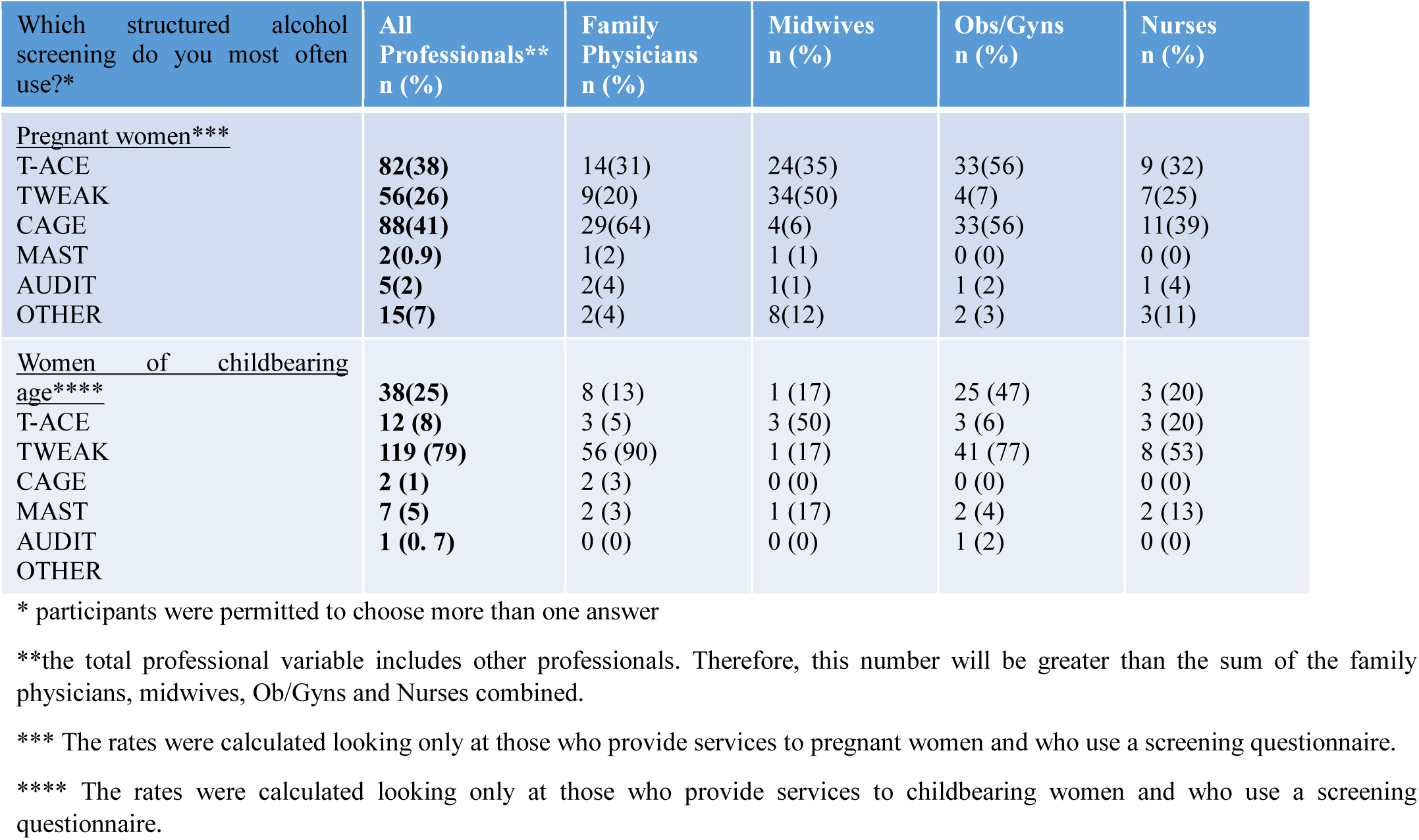
Choice of alcohol screening questionnaire used by clinicians.

In addition to understanding screening practices, it was important to determine how HCPs responded to positive screens. All respondents reported low rates of brief intervention when at-risk drinking was identified in pregnant women (16.4%), and women of childbearing age (39.9%). Family physicians were the most likely to provide brief intervention for both groups of women. The highest rates of referral to harm reduction and treatment programs were found for women of childbearing age when at-risk drinking was identified (50.6%) compared to pregnant women when at-risk drinking was identified (38.7%). Midwives and nurses reported the highest referral rates for pregnant women, whereas family physicians reported the highest referrals rates for women of childbearing age.

## Discussion

This study explored the alcohol screening practices of Canadian women’s HCPs. Although most respondents reported asking about alcohol use, overall a smaller percentage actually reported using validated screening questionnaire. This is consistent with the findings from a descriptive study by Davis and colleagues exploring alcohol risk assessment practices of Canadian primary care professionals (Davis et al., 2008). The majority of respondents in their study reported that they either “rarely or never” used a standardized screening questionnaire *or* that they used a questionnaire that was less sensitive. Practices varied according to gender, length of time in practice, and practice location (Davis et al., 2008). Poole and colleagues reported that screening, BI, and referral to treatment (SBIRT) was not provided universally or systematically across Canada (Poole et al., 2016). Low screening rates may translate into missed opportunities for intervention, resulting in alcohol exposed-pregnancies and suboptimal maternal, neonatal, and child outcomes.

A review of the qualitative data collected in this survey revealed that HCPs, who do not use validated screening questionnaires, identified the following barriers to use: insufficient time; lack of education and/or awareness of screening questionnaires; lack of access to screening questionnaires; and the perception that screening was out of their scope of practice (data not shown). This is consistent with other studies that reported lack of knowledge (Poole et al., 2016) and perceived lack of time and competence in BI (Wangberg, 2015) as barriers to integrating effective screening into clinical practice.

A national survey of Swedish midwives (n=2106) revealed that although they had excellent or good knowledge concerning the risks associated with drinking alcohol during pregnancy, they lacked the skills to identify pregnant women with at-risk pre-pregnancy alcohol use. Resources related to alcohol use during pregnancy for parents, and strategies to manage at-risk drinking behaviours were identified as knowledge gaps (Holmqvist & Nilsen, 2010).

### Strategies to Increase Alcohol Screening Practices

HCPs may benefit from education to improve their use of evidence-based screening questionnaires. It is important for HCPs to understand the evidence underlying efficacy of screening questionnaires and the low time investment, in most cases, needed to implement screening practices in routine care.

Embedding a validated alcohol screening questionnaire into prenatal records may help improve uptake by increasing awareness and ready access for busy HCPs (Premji & Semenic, 2009). Ontario, the province with the greatest representation in this survey, released the newest version of their prenatal record in 2017 and now includes the T-ACE questionnaire (PCMCH, 2017). It is anticipated that alcohol screening practices will improve, as HCPs use this evidence-based questionnaire as part of routine prenatal care provision. Additionally, utilizing standard drink resources can assist patients to quantify their alcohol consumption and improve accurate documentation (Butt 2011; Premji & Semenic, 2009).

When developing and delivering education programs on alcohol use in pregnancy, application-type modules and/or activities are preferable to a solely didactic approach. In one study, HCPs in the prenatal care setting screened 95% of pregnant women for alcohol use after receiving training on the use of screening questionnaires and BI (Kennedy, Finkelstein, Hutchins, & Mahoney, 2004). Important components of the training were: integrating skill building sessions and information delivered by physicians with expertise in this field; identifying community treatment resources and referral pathways; reviewing a BI protocol and clinical decision tree/protocol, as well as offering ongoing technical assistance and consultation (Kennedy et al., 2004). In another study, a one day training session combined with continuous expert support was sufficient for a group of Swedish midwives to implement systematic screening during regular prenatal care visits (Goransson, Magnusson, & Heilig, 2006).

The use of technology could also be further explored for novel mechanisms for the implementation or enhancement of SBIRT in clinical settings. This could be further explored as a cost-effective alternative to traditional counselling approaches (Wilton et al., 2013). Computerized alcohol screening and BI was implemented at a public health clinic to 290 pregnant women with positive outcomes (Nayak, Korcha, Kaskutas, & Avalos, 2014). Eighty-seven percent of participants completed the online program and 91% completed the key screening and BI modules. The program identified alcohol use in 21% of pregnant patients, which was higher than the rates obtained after screening by clinic staff (13%, p<0.01). The majority of program participants (97%) reported that the program was easy to use, they learned something new (88%), and would share what they learned with others (83%) (Nayak et al., 2014).

### Future Directions

Future efforts of Canadian HCP associations and societies should consider the gaps identified in this study and explore opportunities to meet the priority needs. Standardized curricula regarding substance use should be included in all undergraduate and postgraduate health professionals programs, or offered as specialized courses for continuing professional education credit.

At the community level, more referrals to treatment programs are required, especially when at-risk drinking is identified in pregnant women. It is unclear as to why, in the current study, there were fewer referrals to harm reduction and treatment program when women were pregnant. This could reflect a lack of knowledge about available treatment programs, or a lack of programs to which HCPs can refer women. A gap in treatment services has been previously identified in Canada (Poole et al., 2016), as well as, the United States (Diekman et al., 2000). Poole and colleagues acknowledged that while pregnant women are often given priority access to substance use treatment, there is a need for tailored, gender-responsive and pregnancy-focused substance use treatment programs (Poole et al., 2016). Access is another consideration, especially for women living in rural and remote communities. Furthermore, programs that support women’s recovery, while remaining with their babies and children, are needed.

Access to SBIRT in the preconception and interconception period has also been cited as a key facilitator in reducing the risk of alcohol exposed pregnancies (Poole et al., 2016). Preconception interventions have been documented to be effective in reducing at-risk drinking, increasing the use of contraception, or both (Scholin, 2016).

### Limitations

This study has a number of limitations. A high number of respondents were midwives and there was over-representation from Ontario compared to other provinces and territories. While the number of midwives providing care across the country is growing (PHAC, 2012), a significant proportion of early prenatal care is provided by family physicians. Similarly, the low number of responses of HCPs who care for women of childbearing age may be a reflection of the underrepresentation of family physicians and Ob/Gyns who often see these women for other medical reasons.

Further research should explore the predictors of HCPs’ use of alcohol screening questionnaires, in particular, years in practice, practice location, attitudes about screening, and attitudes about BI to address any gaps in knowledge and novel approaches to education and training. These findings would help to target future knowledge translation activities.

## Conclusion

All HCPs caring for pregnant women or women of childbearing age have an essential role in identifying and mitigating problematic alcohol use. There are a number of evidence-based questionnaires that can be used to assess alcohol use. HCPs should strive to provide a safe space to have open, non-judgemental conversations that explore the contributing factors to a woman’s alcohol use and provide BI as indicated. In doing so, comprehensive assessments can be completed and individualized care can be provided.

## Data Availability

Requests regarding data can be sent to the corresponding author.

## Acknowledgements

This research was supported by a Public Health Agency of Canada contribution agreement.

## List of Abbreviations

BI: Brief Interventions
SOGC: Society of Obstetricians and Gynaecologists of Canada
ACOG: American College of Obstetrics and Gynecology
T-ACE: Tolerance, Annoy, Cut down, Eye-opener
TWEAK: Tolerance, Worry, Eye-opener, Amnesia, Kut down
CAGE: Cut down, Annoy, Guilt, Eye-opener
CRAFFT: Car, Relax, Alone, Forget, Family, Trouble
AUDIT-C: Alcohol Use Disorders Identification Test – Alcohol Consumption Questionnaire
SMAST: Shortened Michigan Alcohol Screening Test
RCT: Randomized Controlled Trial
SBIR: Screening, Brief Intervention and Referral
SBIRT: Screening, Brief Intervention, and Referral to Treatment
FASD: Fetal Alcohol Spectrum Disorders

